# Relationship between teaching modality and COVID-19, well-being, and teaching satisfaction (Campus & Corona): a cohort study among students in higher education

**DOI:** 10.1101/2021.03.26.21254388

**Authors:** Atle Fretheim, Arnfinn Helleve, Borghild Løyland, Ida Hellum Sandbekken, Martin Flatø, Kjetil Telle, Sara Sofie Viksmoen Watle, Alexander Schjøll, Sølvi Helseth, Gro Jamtvedt, Rannveig Kaldager Hart

## Abstract

**Background:** After lock-down during the first wave of the COVID-19 pandemic, higher education institutions globally struggled to balance the need for infection control and educational requirements as they prepared to reopen. A particularly difficult choice was whether to offer for in-person or online teaching, since there was little or no empirical research to inform this decision. Norwegian universities and university colleges opted for a hybrid model when they reopened for the autumn semester, with some students offered more in-person teaching than others. This gave us an opportunity to study the association between different teaching modalities and COVID-19 risk, quality of life (subjective well-being), and teaching satisfaction.

**Methods:** We conducted a prospective, observational cohort study among students in higher education institutions in Norway. Participants were surveyed biweekly from September to December in 2020.

**Findings:** 26 754 students from 14 higher education institutions provided data to our analyses. Our best estimate for the association between two weeks of in-person teaching and COVID-19 risk was −22% (95% CI −77% to 33%), compared to online teaching. Quality of life was positively associated with in-person teaching (3% relative risk difference; 95% CI 2% to 4%), as was teaching satisfaction (10%; 95% CI 8% to 11%).

**Interpretation:** The association between COVID-19 infection and teaching modality was highly uncertain. Shifting from in-person to online teaching seems to have a negative impact on the well-being of students in higher education.

**Funding:** None.

## Background

Higher education institutions around the world shut down during the first half of 2020, when the first wave of the COVID-19 pandemic struck. A key question, as institutions prepared to reopen for students, was how teaching could take place without compromising the need for infection control, and specifically whether universities and colleges should switch from in-person to online teaching.^1^

The degree to which different teaching modalities impact on the spread of infection, is uncertain. As Gressman and Peck put it: “In the absence of relevant prior experience, these institutions are largely in the dark about how one might expect a COVID-19 outbreak to evolve in the unique environment of a college campus and how much of an effect the many possible mitigation strategies should be expected to produce.”^2^ Thus, the leadership at higher education institutions faced difficult decisions when trying to weigh the need for infection control against other considerations, e.g. psychosocial, educational and societal consequences.^3-5^

Early on, attempts were made at modelling the risk of offering in-person teaching on campus, e.g. a group at Cornell University arrived at the counter intuitive conclusion that shifting to online teaching would lead to more COVID-19 cases than a full return of students. However, this was premised on “aggressive asymptomatic surveillance where every member of the campus community is tested every 5 days”, as well as sufficient capacity for quarantining, and a series of other assumptions.^6^

Brauner and colleagues exploited the variation in timing of non-pharmaceutical interventions across 41 countries from January to May 2020, to disentangle the impact of individual interventions. They found that concurrent closing of schools and universities was associated with a 38% (95% CI 16% to 54%) reduction of the effective reproductive number.^7^ This is perhaps the most convincing study of non-pharmaceutical interventions to date, despite important potential weaknesses that the authors also acknowledge, e.g. that “the data are retrospective and observational, meaning that unobserved factors could confound the results”, and that “effectiveness estimates can be highly sensitive to arbitrary modelling decisions.”^7^

When universities and colleges reopened after the first pandemic wave, typically in August/September 2020, they opted for different teaching models. Some largely offered in-person teaching, while others mainly offered teaching online.^8^ This variation has been utilized in a handful of studies from the United States, by linking information on teaching modalities with county level data on COVID-19 incidence.^9-12^ These studies seem to show that transmission has been higher in counties hosting institutions that have offered in-person teaching than in counties where institutions mainly offered online teaching. However, consistent findings across these studies is not surprising as they are largely based on the same data. Whereas these studies have shown associations using county-level data, this is the first study to our knowledge of the individual impacts of shifting from in-person to online teaching in higher education. Apart from these American studies, remarkably little empirical work has been done to assess the consequences of shifting from in-person to online teaching in higher education as an infection control measure.

While the evidence base is weak for shifting from in-person to online teaching to control the spread of COVID-19, it is even weaker for other outcomes, such as well-being and quality of life. A few cross sectional surveys of students in higher education have reported on the prevalence of psychological outcomes or quality of life measures during the COVID-19 pandemic, but to our knowledge none have compared different teaching modalities.^13-16^

In Norway, higher education institutions opted for a hybrid model when they reopened for the autumn semester in August 2020, by offering more in-person teaching to some students and more online teaching to others. For instance, many institutions prioritised first year students for in-person teaching. The variation across students and over time in teaching modality gave us an opportunity to study the relationship between exposure to in-person versus online teaching, and key outcomes.

Our aim with this study was to assess the association between teaching modality (in-person or online teaching), and COVID-19, well-being, and satisfaction with teaching, among higher education students in Norway.

## Methods

Our reporting adheres to the STROBE-statement. We registered the study in advance at ClinicalTrials.gov (Identifier: NCT04529421) and published the study protocol with analysis plan before recruitment started.^17^

The study took place from September to December 2020. It was initiated from Oslo Metropolitan University and invitations to participate were sent from the university’s rector to 32 universities and university colleges in Norway. The Institutions that agreed to take part in the study granted us access to their students’ contact information (e-mail addresses and telephone numbers).

All students who were registered at the participating institutions were invited by SMS and/or e-mail. The invitation included a link to a web-based informed consent form and a questionnaire. We sent two reminders. Students who consented, received a new SMS/e-mail with link to the questionnaire every two weeks during the study period, i.e. up to 8 times. The consent form and questionnaire were available in both Norwegian and English.

We developed the questionnaire through an iterative process, partly based on items from existing questionnaires. Pilot testing was done with a group of 10 students at Oslo Metropolitan University. The questionnaire took less than 10 minutes to complete.

The questionnaire contained items on how much in-person and online teaching they had been offered over the last two weeks, and how much time they had spent on campus. We also inquired about testing for COVID-19, subjective well-being, satisfaction with teaching, social activities, and more. See Supplement 1 for full questionnaire.

The participants consented to linking their survey data to the administrative data system for higher education in Norway, the Common Student System, used by all higher education institutions in the study. This gave us access to information about which study program each student was enrolled in, basis for admission, study status, academic results, gender, and age (see Supplement 2 for full list of variables).

During the study period we realised that some groups of students were of limited relevance to our study, i.e. master-level students who spend most of their time writing their theses, and post-graduate students who typically spend a small part of their time in class. We therefore limited our analyses to students registered as 1^st^, 2^nd^, or 3^rd^ year bachelor level students. This decision was made before data analysis.

### Primary outcome

- COVID-19 incidence (self-reported positive test results)

### Secondary outcomes

- Well-being (“Overall, how satisfied are you with life right now?” on a 0-10 scale)
- Teaching satisfaction (“Overall, how satisfied have you been with the teaching you have received in the past 14 days?” on a 0-10 scale)
- COVID-19 testing (self-reported)
- Quarantine (self-reported)

We added the question about quarantine after we published the protocol. Data on learning outcomes are not yet available and will be reported at a later stage.

We defined in-person teaching as a continuous variable:

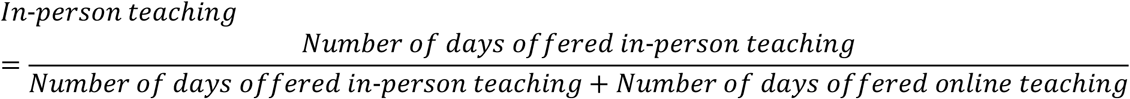

### Analytical approach

We ran multivariate regressions (ordinary least squares) to test for associations between in-person teaching and the outcomes. We adjusted for a range of potentially confounding variables, both time constant (institution, year and field of study (and their interaction) parents’ country of origin, own country of origin, gender, age, age squared, parents’ educational level) and time varying (number of roommates, home ownership, total proportion in quarantine at institution, alcohol consumption, use of public transport, total amount of offered teaching, and number of hours of paid work).

We decided, during the analysis phase that we needed to account for coinciding infection control measures. To this end, we included the proportion of study participants (excluding the individual) at the same institution who were in quarantine, as a proxy control variable.

We collected exposure and outcome data from the same response questionnaire. It is therefore possible that the outcome (positive COVID-19 test) happened before the exposure (teaching modality) in some cases.

Finally, we also ran analyses using self-reported actual on campus presence as the independent variable.

### Sensitivity analyses

We conducted the following sensitivity analyses to check the robustness of our main findings:

- Comparing the quartile with most in-person teaching to the quartile with least
- Changing the exposure variable from continuous (proportion of in-person teaching) to dichotomous (>80 % in-person teaching, or not)
- Restricting data for COVID-19 related outcomes to data from the period immediately following the period when data on teaching modality was collected (lag model)
- Replacing proportion of students in quarantine with an interaction between institution dummies and survey round dummies, netting out institution-specific time shocks (such as a sudden COVID-19 outbreak at one institution)
- Using logit and negative binomial models to check the robustness of our findings for positive COVID-19 tests, testing, and quarantining.

As a robustness check we also ran a lead model, i.e. we tested for associations between data on outcomes preceding data on teaching modality. A statistically significant (p<0.05) association between outcomes and future predictors would indicate that confounders are present.

Recognising the potential importance of coinciding infection control measures as confounding variables, we decided to explore whether controlling for county level COVID-19 incidence in the week preceding each survey round would influence our findings.

We also decided to conduct a fixed effects analysis, i.e. assessing the association between teaching modality and outcomes based on the variation we observed for each individual participant, instead of comparing across participants. The main advantage with the fixed effects approach is that it accounts for measured and unmeasured differences between participants, since all comparisons are based on data for the same participant at different points in time, not between different participants. A limitation of the fixed effect approach is that we only capture effects of changes in teaching modality that individual students are exposed to, which also means that we can only include data from participants who have responded to the questionnaire at least twice.

### Ethics and data protection

We used University of Oslo’s solutions for online consent form, web-based survey (‘Nettskjema’), and secure storage of research data (TSD).

Data management was in accordance with GDPR-regulations. Our Data Protection Officer at the Norwegian Centre for Research Data assessed the project plan and found it satisfactory.

The Regional Ethical Committee assessed and approved the project plan (24 August, REK sør-øst A, reference number 172155).

### Power analysis and sample size

We based our power calculation on the assumption that 0.23% (230 per 100.000) of the participants with predominantly online teaching would report testing positive for COVID-19, over a 10-week intervention period – corresponding to the incidence for the age group 20-29 in Norway at the time. To detect an effect of in-person teaching corresponding to a doubling of COVID-19 risk, we estimated a need for 21 000 respondents to be 80% certain to detect an effect at the 5% significance level.

## Results

The leadership at 14 universities and university colleges, with around 45% of all students in higher education in Norway, agreed to take part in the study.^18^

We invited all 142 384 registered students at the 14 institutions. 35 423 of these students responded to the survey at least once. We excluded part-time students and students that were not 1^st^, 2^nd^, or 3^rd^ year students. This left us with 26 754 participants that we included in our analyses. The number of responses per survey round is shown in Figure 1. The total number of responses was 72 369, meaning that we collected around 5 ½ weeks’ worth of data per participant, on average.

**Figure 1:**
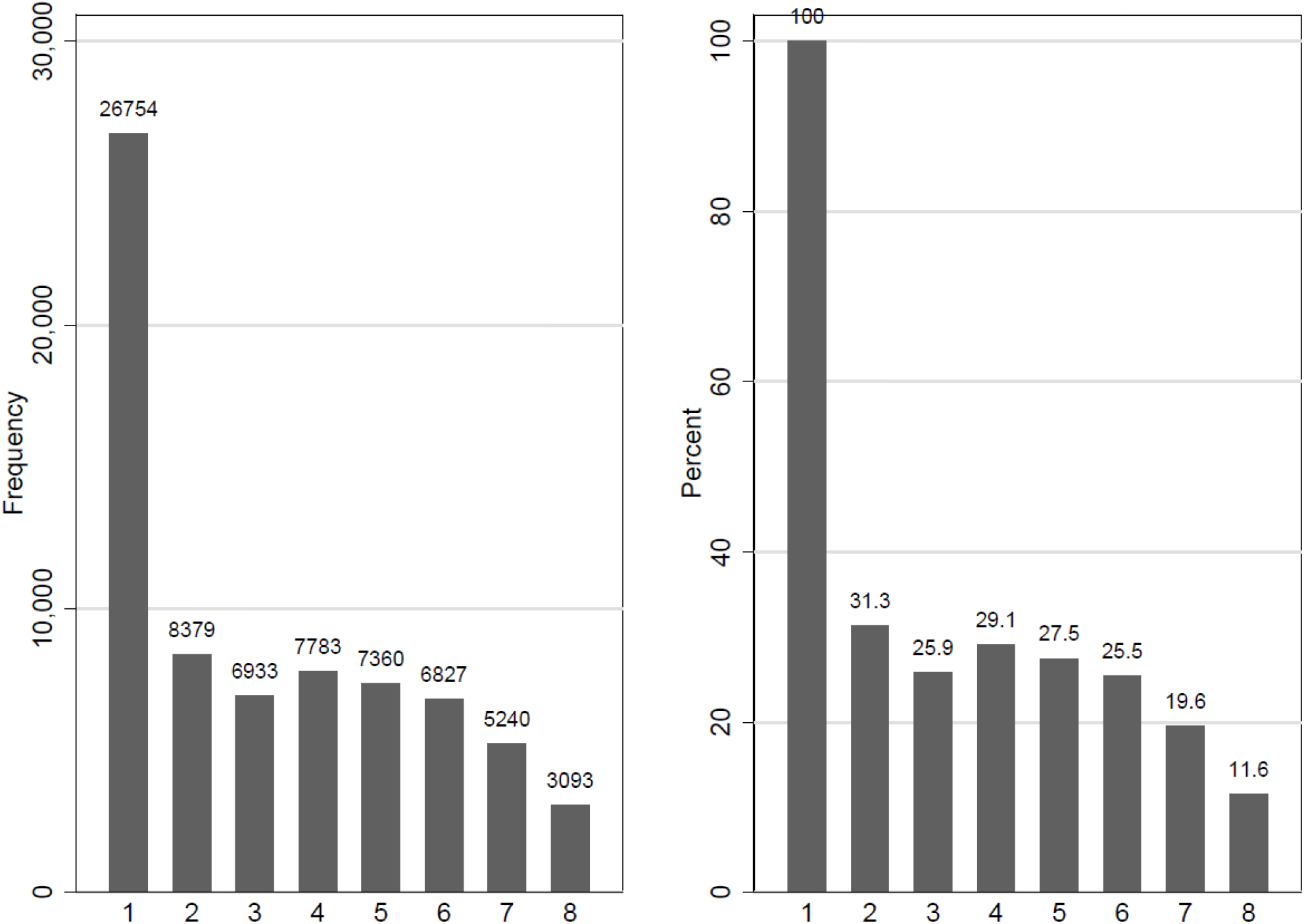
Attrition and number of respondents over survey rounds.

Figure 2 is a presentation of how teaching modality and outcomes changed during the study period. Over the semester, there was a marked fall in the proportion in-person teaching. Well-being and satisfaction with teaching also fell throughout the semester, with a small increase in well-being towards the end of the semester, before the Christmas holidays. In general, there were more positive COVID-19 tests, more testing and more quarantine in the latter part of the semester than in the earlier.

**Figure 2:**
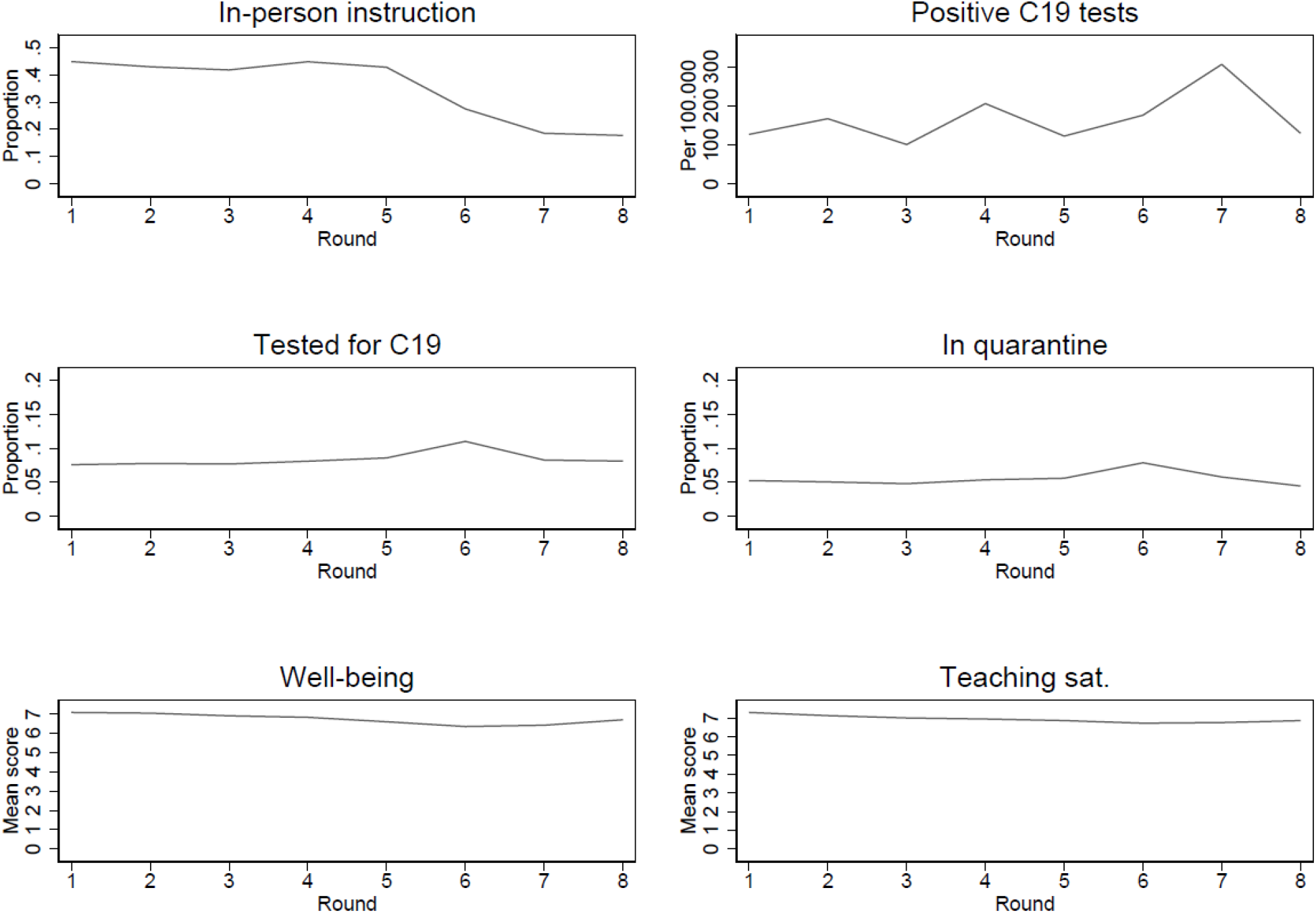
Trends in main outcomes over survey rounds.

Key background variables are shown in Table 1, where we have divided the participants into quartiles according to the proportion of in-person teaching they were offered. Results from balancing tests are shown in Table S1. The difference across modality quartiles was statistically significant (p<0.05) for gender, parental education, and migration background, but became insignificant after controls for year and field of study were added. For age, this difference persisted also when all the association was purged of other observed covariates.

**Table 1:** Descriptive statistics for background variables

|  |  |  | By quartile of proportion in-person teaching |  |  |  |
| --- | --- | --- | --- | --- | --- | --- |
| Variable | Category | All | Q1 | Q2 | Q3 | Q4 |
| Male | No | 18 244<br>(68·2%) | 3404<br>(70·7%) | 4939<br>(71·5%) | 4896<br>(66·6%) | 5005<br>(65·1%) |
|  | Yes | 8510<br>(31·8%) | 1412<br>(29·3%) | 1968<br>(28·5%) | 2451<br>(33·4%) | 2679<br>(34·9%) |
| Parents higher education | No | 8070<br>(30·2%) | 1444<br>(30·0%) | 2115<br>(30·6%) | 2234<br>(30·4%) | 2277<br>(29·6%) |
|  | Yes | 18684<br>(69·8%) | 3372<br>(70·0%) | 4792<br>(69·4%) | 5113<br>(69·6%) | 5407<br>(70·4%) |
| Respondents' place of birth | Norway | 22811<br>(85·3%) | 3930<br>(81·6%) | 5956<br>(86·2%) | 6285<br>(85·5%) | 6640<br>(86·4%) |
|  | Europe | 1774<br>(6·6%) | 374<br>(7·8%) | 398<br>(5·8%) | 489<br>(6·7%) | 513<br>(6·7%) |
|  | Africa | 422<br>(1·6%) | 109<br>(2·3%) | 113<br>(1·6%) | 107<br>(1·5%) | 93 (1·2%) |
|  | Asia | 1338<br>(5·0%) | 290<br>(6·0%) | 357<br>(5·2%) | 359<br>(4·9%) | 332<br>(4·3%) |
|  | Australia / Oceania | 17 (0·1%) | 6 (0·1%) | 5 (0·1%) | 2 (<1%) | 4 (0·1%) |
|  | North America | 185<br>(0·7%) | 53 (1·1%) | 31 (0·4%) | 52<br>(0·7%) | 49 (0·6%) |
|  | South or Central America | 165<br>(0·6%) | 47 (1·0%) | 41 (0·6%) | 35<br>(0·5%) | 42 (0·5%) |
|  | Not reported | 42 (0·2%) | 7 (0·1%) | 6 (0·1%) | 18<br>(0·2%) | 11 (0·1%) |
| Parents' place of birth | Norway | 20177<br>(75·4%) | 3504<br>(72·8%) | 5210<br>(75·4%) | 5540<br>(75·4%) | 5923<br>(77·1%) |
|  | Europe | 3036<br>(11·3%) | 580<br>(12·0%) | 721<br>(10·4%) | 850<br>(11·6%) | 885<br>(11·5%) |
|  | Africa | 670<br>(2·5%) | 159<br>(3·3%) | 191<br>(2·8%) | 181<br>(2·5%) | 139<br>(1·8%) |
|  | Asia | 1922<br>(7·2%) | 307<br>(6·4%) | 571<br>(8·3%) | 545<br>(7·4%) | 499<br>(6·5%) |
|  | Australia / Oceania | 35 (0·1%) | 11 (0·2%) | 10 (0·1%) | 5 (0·1%) | 9 (0·1%) |
|  | North America | 325<br>(1·2%) | 74 (1·5%) | 66 (1·0%) | 87<br>(1·2%) | 98 (1·3%) |
|  | South or Central America | 253<br>(0·9%) | 61 (1·3%) | 74 (1·1%) | 62<br>(0·8%) | 56 (0·7%) |
|  | Not reported | 336<br>(1·3%) | 7 (0·1%) | 6 (0·1%) | 18<br>(0·2%) | 11 (0·1%) |
| Age (SD) |  | 25·14<br>(6·84) | 26·56<br>(7·63) | 24·61<br>(6·36) | 24·94<br>(6·83) | 24·91<br>(6·64) |
| Year of study (SD) |  | 1·61<br>(0·76) | 1·76<br>(0·80) | 1·63<br>(0·75) | 1·55<br>(0·73) | 1·56<br>(0·74) |
| Previous positive<br>COVID-19 test (SD) |  | 127·1<br>(3562·7) | 207·6<br>(4552·5) | 144·8<br>(3802·5) | 81·7<br>(2586·8) | 104·1<br>(3225·2) |
| Hours of paid work last<br>14 days (SD) |  | 14·6<br>(18·4) | 17·5<br>(20·9) | 14·8<br>(17·7) | 14·0<br>(18·0) | 13·2<br>(17·6) |
| N |  | 26 754 | 4816 | 6907 | 7347 | 7684 |

Table 2 shows outcomes measured throughout the observation period. A total of 112 participants reported having had a positive COVID-19 tests. We found the highest incidence of COVID-19 in the quartile with least in-person teaching offered (206 cases per 100 000), and the lowest incidence in the quartile with most in-person teaching (123 per 100 000). Quarantining and testing followed the same pattern, with 9% and 6% being tested and quarantined respectively, in the quartile offered least in-person teaching. For the quartile offered most in-person teaching, the corresponding figures were 7% and 5%, respectively. Well-being and teaching satisfaction were lowest in the quartile with least in-person teaching (6.5 on a scale from 0 to 10 for both outcomes), and highest in the top quartile (7.2 and 7.8 for teaching satisfaction and well-being, respectively).

**Table 2:** Descriptive statistics for outcome variables. Means and standard deviations.

| Variable | All | By quartile of proportion in-person teaching |  |  |  |
| --- | --- | --- | --- | --- | --- |
|  |  | Q1 | Q2 | Q3 | Q4 |
| Positive test (per 100 000) | 155.0<br>(3934.4) | 206.3<br>(4536.9) | 149.8<br>(3867.8) | 134.6<br>(3666.8) | 123.3<br>(3505.6) |
| Tested | 0.08 (0.27) | 0.09 (0.29) | 0.09 (0.29) | 0.08 (0.26) | 0.07 (0.26) |
| Quarantined | 0.05 (0.23) | 0.06 (0.24) | 0.06 (0.24) | 0.05 (0.22) | 0.05 (0.21) |
| Satisfied with teaching | 7.06 (2.52) | 6.53 (2.58) | 6.55 (2.44) | 7.35 (2.30) | 7.83 (2.49) |
| Well-being | 6.85 (2.33) | 6.54 (2.40) | 6.66 (2.32) | 7.00 (2.27) | 7.21 (2.27) |
| N | 72 243 | 19 879 | 16 687 | 17 827 | 17 850 |

### Main findings

Our main findings are presented in Table 3.

**Table 3:** Main results, coefficients (95% confidence intervals) from ordinary least squares analyses. Relative scale is obtained by dividing the coefficient and their standard error on the average of the outcome when the explanatory variable takes zero.

|  | Absolute scale |  | Relative scale |  |
| --- | --- | --- | --- | --- |
|  | Unadjusted | Adjusted | Unadjusted | Adjusted |
| **COVID-19 positive** |  |  |  |  |
| Proportion in-person | -91.0* | -49.6 | -41.2* | -22.4 |
|  | (-171.6 to -10.4) | (-171.0 to 71.8) | (-77.7 to -4.7) | (-77.4 to 32.5) |
| N | 72 243 | 69 930 | 72 243 | 69 930 |
| **COVID-19 tested** |  |  |  |  |
| Proportion in-person | 0.0*** | 0.0 | -25.7*** | -3.7 |
|  | (0.0 to 0.0) | (0.0 to 0.0) | (-32.4 to -19.0) | (-13.2 to 5.8) |
| N | 72 369 | 70 055 | 72 369 | 70 055 |
| **Quarantine** |  |  |  |  |
| Proportion in-person | 0.0*** | 0.0 | -28.1*** | -11.0 |
|  | (0.0 to 0.0) | (0.0 to 0.0) | (-36.5 to -19.6) | (-23.0 to 1.1) |
| N | 72 369 | 70 055 | 72 369 | 70 055 |
| **Satisfied with teaching** |  |  |  |  |
| Proportion in-person | 1.6*** | 1.0*** | 24.4*** | 15.7*** |
|  | (1.5 to 1.7) | (0.9 to 1.1) | (23.2 to 25.6) | (14.3 to 17.1) |
| N | 71 679 | 69 391 | 71 679 | 69 391 |
| **Well-being** |  |  |  |  |
| Proportion in-person | 0.8*** | 0.4*** | 12.1*** | 6.8*** |
|  | (0.7 to 0.9) | (0.4 to 0.5) | (11.0 to 13.2) | (5.5 to 8.1) |
| N | 72 110 | 69 839 | 72 110 | 69 839 |

\*p<0.05, \*\*p< 0.01, \*\*\*p<0.001. Note: Adjusted model includes controls for institution, year and field of study (and their interaction) parents' country of origin, own country of origin, gender, age, age squared, parents' educational level, number of roommates, home ownership, total proportion in quarantine at institution, alcohol consumption, use of public transport, total amount of offered teaching, and number of hours of paid work.

The unadjusted bivariate analysis yielded a negative association corresponding to a 41% reduction in the probability of COVID-19 (95% CI −78% to −5%) for two weeks of full time in-person teaching compared to full time online teaching. In the adjusted analysis, the association between in-person teaching and positive COVID-19 test remained negative, but was smaller and statistically non-significant (22% reduction, 95% CI −77% to 33%, see Table 3).

The findings were similar for the other COVID-19 related outcomes, but the associations were weaker: In the unadjusted analyses, the associations for COVID-19 testing and quarantine were −26% (95% CI −32 to −19%), and −28% (95% CI −37% to −20%) respectively, for two weeks of full time in-person teaching compared to full time online teaching (see Table 3). In the adjusted analyses the corresponding associations were reduced to −4% (95% CI −13% to 6%) and to −11% (95% CI −23% to 1%).

For well-being and teaching satisfaction we found positive associations with in-person teaching in the unadjusted analyses, and the results remained statistically significant in the adjusted analyses: Without controlling, two weeks of in-person teaching was associated with a 12% (95% CI 11% to 13%) higher well-being and a 24% (95% CI 23% to 26%) higher teaching satisfaction, relative to two weeks of online teaching. In the adjusted analyses, the positive associations with in-person teaching were 7% for well-being (95% CI 6% to 8%) and 16% for satisfaction with teaching (95% CI 14% to 17%).

When we used actual campus presence as the independent variable instead of offered teaching modality, we found associations of comparable magnitude for well-being and teaching satisfaction (Figure S1, Panel B). COVID-19 positive test, COVID-19 testing, and quarantine were all negatively associated with campus presence. This was as expected, since those with COVID-19 symptoms or in quarantine had stricter social distancing rules to adhere to.

### Sensitivity analyses

The comparison between the quartiles with most and least in-person teaching (Figure S2, Panel A), and the comparison of students with 80% or more in-person teaching against all other students (Figure S2, Panel B), yielded results that were largely in line with our main findings.

The results for COVID-19 related outcomes were robust to lagging the outcome with one round (Figure S3).

Neither replacing the proportion of students in quarantine with institution specific trends as a control variable, or controlling for county level COVID-19 incidences, or controlling for paid working time, led to substantially different results (data not shown).

The results from the lead model estimated by ordinary least squares (Figure S1, Panel C) showed no significant effects for positive COVID-19 test, testing or quarantine, while there were significant lead effects for well-being and teaching satisfaction.

When we applied the fixed effects approach, the findings were largely compatible with the results from our main model (Table S2 and Figure S1, Panel D). All estimates for positive COVID-19 tests were substantially larger, but they remained statistically non-significant (p<0·05). For quarantine and testing, the fixed effect approach yielded similar estimates as the main analyses. The estimates for well-being and teaching satisfaction were nearly halved in the fixed effects analyses, but they remained statistically significant. The difference between our main results and the fixed effect analyses, suggests that individuals more satisfied with life and with teaching tended to be in programs offering more in-person teaching.

We found no statistically significant lead effects for the fixed effects models (Figure S1, Panel E).

Results from running our data through logit and negative binomial models were consistent with our main findings (data not shown).

## Discussion

Our findings do not demonstrate a convincing association between teaching modality and COVID-19 risk among students in higher education. While the point estimate goes in the direction of a negative association between in-person teaching and COVID-19 risk, the lack of precision in our results means that we cannot rule out important effects in either direction.

However, we did find a relatively convincing positive association between in-person teaching and well-being, and between in-person teaching and teaching satisfaction. These findings are substantiated through a set of sensitivity analyses and do, in our judgement, provide evidence of downsides associated with shifting from in-person to online teaching. Still, the observational nature of our study means that we cannot ignore the risk that confounding may have biased the results.

Although the estimate for the association between in-person teaching and COVID-19 risk is highly uncertain, the negative direction of the association surprised us. Attempting to explain this, we hypothesised that online teaching may lead to increased extracurricular activities due to unmet social needs, and that these activities may increase the risk of COVID-19. We therefore estimated the association between in-person teaching and social activities based on responses to the question, “In the past 14 days, have you been to a social gathering where you would guess that there were 20 or more people?” The findings provided no support for our hypothesis: In-person teaching was associated with more, not less, social activities than was online teaching. This finding may suggest, however, that a shift to more in-person teaching is correlated with fewer social distancing interventions and behaviour in other domains, and that reductions in restrictions may have contributed to the effect on students’ improved well-being. Our attempts to adjust for this, e.g. by including the proportion of students in quarantine or the incidence of COVID-19 in the area in the model, may not have controlled sufficiently for confounding due to changes in restrictions. For teaching satisfaction, we believe it is unlikely that changes in social activity restrictions had an impact.

We employed two different analytical methods, i.e. ordinary least squares and fixed-effect multivariate regression. The ordinary least squares approach served as the primary analysis, in line with our study protocol. However, there are good reasons for putting more emphasis on the findings from the fixed effects approach, at least for some outcomes. For the COVID-19 related outcomes, the difference between the results from the two methods is of limited interest, since they both yielded highly uncertain estimates with wide confidence intervals that bar us from drawing meaningful conclusions. The case is different for the two non-COVID-19 outcomes, well-being and teaching satisfaction. Here, both the ordinary least squares and fixed effects approaches yielded statistically significant positive associations with in-person teaching, but the estimates were substantially smaller in the fixed-effect model. We believe the fixed-effects model provides more credible estimates of the causal effect for these outcomes, since for the ordinary least squares model there were positive associations with in-person teaching also when outcomes came first (lead model). This was not the case for the fixed-effects model; however, this does not exclude the possibility that associations may be driven by correlations between exposures and outcomes at different time points (i.e. autocorrelation). The estimates from the fixed effects models were also the more conservative. Thus, a reasonable interpretation is that full time in-person teaching for two weeks was associated with a relative increase in well-being of 3% (95% CI 2% to 4%), compared to full time online teaching, i.e. a difference of around 0·2 on the 0 to 10-scale used in the study. Correspondingly, our best estimate for teaching satisfaction was a 10% increase (95% CI 8% to 11%).

Our estimates are based on two-week periods. The relationship between teaching modality and outcomes may be expected to increase over time, in which case our estimate for the relationship between online teaching and well-being are underestimates if the teaching period goes beyond two weeks.

This may be the most rigorous study assessing the impact of a large-scale social distancing intervention so far, in the COVID-19 pandemic. We are only aware of one randomised study in this area – the TRAiN-trial, where members of training centres were individually randomised to regain access to their gyms during national lock down.^19^ Also, we are not aware of other prospective observational studies using individual level data, than ours. Still, we were not able to generate conclusive results for the relationship between in-person teaching and COVID-19 risk. There are several reasons for this, most notably that the COVID-19 incidence was substantially lower than we expected, both in in Norway generally, and among the participants specifically. Thus, there were too few COVID-19 cases to demonstrate even substantial effects with statistical certainty.

It may be considered a weakness that we used a single question to assess the participants’ well-being. While a single item cannot possibly capture all aspects of quality of life, the question we used is recommended for measuring subjective well-being – a key component of quality of life.^20,21^

The few surveys we are aware of that have assessed well-being among higher education students during the COVID-19 pandemic have not explored the impact of different teaching modalities.^14-16^ Rather, they have tried to assess whether using digital platforms or previous experience with online learning have mitigated the impact of societal lock downs on students’ quality of life.^14-16^

Findings from the Norwegian Student Survey which took place while we conducted our study, agree with ours with regard to teaching satisfaction: 72 percent of the survey respondents believed their learning outcome would be better if they could be physically present on campus.^22^

The critical lack of randomised trials in this area has been pointed out before.^23,24^ If groups of students at all higher education institutions in Norway had been randomly allocated to different teaching modalities we would have reduced the risk of bias due to confounding. In addition, we could have sourced outcome data from the Norwegian Emergency Preparedness Register for COVID-19, making us less reliant on self-reporting to assess COVID-19 incidence among study participants. A key barrier for such a study is the legal requirement in the Norwegian Health Research Act to obtain written informed consent from all who participate in health research. In practice, the demand for individual consent from all who are affected makes it impossible to carry out comparative studies where teaching institutions, municipalities, workplaces etc. are allocated to different forms of infection control measures.^25^

The impact of interventions to reduce COVID-19 incidence may vary across time and place, e.g. depending on what other measures have been implemented or which virus variant is the dominant one. Still, we believe that our finding indicating a negative effect on students’ well-being when shifting from in-person to online teaching, is likely to be valid beyond our specific study setting.

## Conclusion

We did not find clear evidence of an association between COVID-19 infection and teaching modality for students in higher education, but our findings indicate that shifting from in-person to online teaching may impact negatively on the students’ well-being.

## Data Availability

After ensuring that re-identification is not possible, the dataset will be stored and made publicly available for at least 10 years at the Norwegian Centre for Research Data (NSD). We aim to have this in place by July 2021. Analytic codes are available upon request to the study chief analyst (RKH).

## Author contributions

AF conceived and initiated the study, contributed in all parts of the project as primary investigator, and wrote the first draft of the manuscript. AH contributed to the planning and management of the project, to developing the questionnaire, and had the lead role in managing the survey. RKH managed the data, conducted all analyses and contributed to the data analysis design. BL and IHS contributed to the development of the questionnaire and conducted the pilot testing of it. AS contributed to the data analysis plan and to developing the questionnaire. SSVW contributed in developing the questionnaire. MF contributed to the questionnaire development, the data analysis plan, and to the analyses. KT played the lead role in writing the data analysis plan and contributed to the analyses. GJ and SH contributed to the planning and designing the study. All authors read, commented on, and approved the final study report.

## Acknowledgements

We thank Ole Petter Ottersen (Karolinska Institute) for motiving the development of this study. We also thank Per Martin Norheim-Martinsen (Oslo Metropolitan University) for supporting the project team in the development phase. The following students at Oslo Metropolitan University tested and gave us feed-back on the draft questionnaire: Rikard Dotzler, Kamilla Frøysland,Åsne Dahl, Tonje Elin Vik, Amina Shahzad, Ola Gimse Estenstad, Karl Aksel Lyby, Ole Jørgen Knoph, Eirin Lindtner Storesund, and Ane Kollen Evenmo. Ann Oldervoll translated the questionnaire and the consent form to English, and Helen Ghebremedhin translated the questionnaire back to Norwegian for quality control (both Norwegian Institute of Public Health). Geir Vangen at the Directorate for ICT and joint services in higher education and research (UNIT) provided administrative data from the Common Student System. Dagfinn Bergsager (University of Oslo) facilitated our use of the web-based questionnaire and safe data storage. For data storage and analyses we used the TSD (Tjeneste for Sensitive Data) facilities, owned by the University of Oslo, operated and developed by the TSD service group at the University of Oslo, IT-Department (USIT). We thank the leadership at the following institutions who facilitated the recruitment of students to the study: Oslo Metropolitan University, University of South-Eastern Norway, University of Oslo, Western Norway University of Applied Sciences, University of Agder, The Arctic University of Norway, Nord University, Lovisenberg Diaconal University College, Norwegian University of Life Sciences, Kristiania University College, Østfold University College, The Oslo School of Architecture and Design, The Norwegian Academy of Music, and Norwegian School of Theology, Religion and Society. Finally, we wish to thank each of the 35 423 students who agreed to participate in this study.

