## Supplemental materials for "Relationship between teaching modality and COVID-19, well-being, and teaching satisfaction (Campus & Corona): a cohort study among students in higher education"

### SUPPLEMENT 1. QUESTIONNAIRE

**How old are you?**

Select 19 or younger

Etc.

50 or older

**With which gender do you identify?**

Woman

Man

Other

Prefer not to answer

**Where were you born?**

Norway

Europe (country other than Norway)

Africa

Asia

Australia/Oceania

North America

South or Central America

**Where were your parents/guardians born?**

(Select all that apply)

Norway

Europe (country other than Norway)

Africa

Asia

Australia/Oceania

North America

South or Central America

**What is your parent's/guardian's highest level of completed education?**

Primary/elementary school ( ) Secondary or high school ( ) College/university ( )

Parent/guardian 1

Parent/guardian 2

**Have you been tested for the corona virus previously this year, before the beginning of the fall semester 2020?**

Yes

No

**In the past 14 days, have you been tested for the corona virus?**

Yes [the Norwegian version used No then yes this time, but the responses should always be in the same order so I am writing yes then no!]

No

**To what extent are you worried about being infected by the corona virus?**

To a very little extent

To a small extent

To a moderate extent

To a great extent

To a very great extent

**In the past 14 days, have you been to a social gathering where you would guess that there were 20 or more people?**

Yes

No

Do not know

**How many times have you participated in-person at a social gathering with your "faddergruppe" (buddy group) this fall semester 2020?**

None

1 time

2 – 4 times

5 times or more

**In the past 14 days, how many times have you drunk 6 or more alcoholic drinks?**

None

1 time

2 times

3 times

4 times

5 times or more

**Which faculty are you enrolled in?**

Faculty of Health Sciences

Faculty of Education and International Studies

Faculty of Social Sciences

Faculty of Technology, Art and Design

**Which department are you affiliated with?**

[Example, if respondent selected Faculty of Technology, Art and Design on previous page]

Department of Civil Engineering and Energy Technology

Department of Mechanical, Electronic and Chemical Engineering

Department of Computer Science

Department of Product Design

Department of Art, Design and Dram

Other affiliation

**For which academic year are you taking most of your courses this semester?**

1st year bachelor's or 1st year integrated master's

2nd year bachelor's or 2nd year integrated master's

3rd year bachelor's or 3rd year integrated master's

1st year master's or 4th year integrated master's

2nd year master's or 5th year integrated master's

Other

I am not studying this semester

**Overall, how satisfied have you been with the teaching you have received in the past 14 days?**

Scale from 0 to 10, where "0" is not satisfied at all and "10" is very satisfied

0 (not satisfied at all)

…

5 (neither satisfied nor dissatisfied)

…

10 (very satisfied)

Do not know

**In the past 14 days, approximately how many days have you been offered in-person instruction (on-campus or elsewhere)?**

Fill in the number of days ____

**In the past 14 days, approximately how many days have you been offered digital (online) instruction?**

Fill in the number of days ____

**In the past 14 days, approximately how many days have you spent on campus (classes, self-study, colloquia and social)?**

Fill in the number of days ____

Have you been able to choose for yourself whether you wanted in-person or digital instruction?

Yes, to a large extent

Yes, to a small extent

No

Do not know

**In the past 14 days, how many days of off-campus work placement have you been offered?**

Fill in the number of days ____

**How many days have you had of off-campus work placement during the past 14 days?**

Fill in the number of days ____

**Approximately how many hours do you work (at a paying job) while studying per week?**

Fill in the number of hours ____

**Who do you live with now?**

Select all that apply

I live alone

With a romantic partner/spouse

With friend(s)/roommate(s)

With parents/guardians

With children

**Who owns the housing unit you live in?**

The welfare organization for students (SiO)

Professional private landlord

Other private landlord

My parents/guardians/relatives

I/we own it

Other

**How many other people do you live with? ____**

**Approximately how many times have you travelled by public transportation (bus, tram, train, boat or plane) in the past 14 days? ___**

**Overall, how satisfied are you with life right now?**

Scale from 0 to 10, where "0" is not satisfied at all and "10" is very satisfied

0 (not satisfied at all)

…

5 (neither satisfied nor dissatisfied)

…

10 (very satisfied)

Do not know

### SUPPLEMENT 2. VARIABLES FROM THE COMMON STUDENT SYSTEM (FS)

1. Sex (M, F)
2. Age
3. Basis of admission
4. Study progression: Average ECTS-credits the last 2 or 3 semesters
5. Grade Point Average (raw)
6. Year of secondary diploma
7. Grade Point Average (“school points”)
8. Grade Point Average (“admission points”)
9. Study programme code
10. Study programme name
11. Study level code
12. Field of study code
13. Field of study name
14. Share of full-time studies
15. Share professional training
16. NUS code
17. Students in study programme
18. Year of study
19. Course name (spring and autumn term 2020)
20. Course code (spring and autumn term 2020)
21. Course level (spring and autumn term 2020)
22. Course completion (spring and autumn term 2020)
23. Planned ECTS-credits (spring and autumn term 2020)
24. Outgoing exchange student (spring and autumn term 2020)
25. Incoming exchange student (spring and autumn term 2020)
26. Course grade (spring and autumn term 2020)
27. Average grade, all courses at institution
28. Institution number (NSD)
29. Institution number (FS)
30. Institution name
31. Place of study code DBH (NSD)
32. Place of study name
33. Campus
34. Faculty name

### SUPPLEMENT 3. SENSITIVITY ANALYSES – RESULTS

### Supplementary Table S1: Results from balancing tests on background variables

| | Balancing tests | | | | |
|---|---|---|---|---|---|
| | **Level** | **Mean** | **Basic** | **Controls*** | **Interactions†** |
| Male | No | *** | *** | N.s. | N.s. |
| Parents higher education | No | N.s. | N.s. | N.s. | N.s. |
| Parents Norwegian born | No | *** | N.s. | N.s. | N.s. |
| Respondent Norwegian born | No | *** | N.s. | N.s. | N.s. |
| Age | | *** | N.s. | *** | *** |
| Year of study | | *** | | | |
| Prev. C19 pos. | | | N.s. | N.s. | N.s. |
| N | 26 401 | 26 401 | 26 401 | 26 401 | 26 401 |

*Controls for proportion in-person teaching, institution, year and field of study. parents' country of origin, own country of origin, gender, age, age squared, parents' educational level, number of roommates, home ownership, and total proportion in quarantine at institution (first survey round), excluding the outcome in question for each row. †Additional controls for the interaction between year and field of study. N.s. = not statistically significant (p<0·0.05). *** <p 0.001. N.s. = Not statistically significant (p>0·0.05).

Supplementary Table S2. Results from individual fixed effects models. Except for individual fixed effects, the models are identical to the main zero. The adjusted model includes controls for institution, year and field of study (and their interaction) parents' country of origin, own country of origin, gender, age, age squared, parents' educational level, number of roommates, home ownership, total proportion in quarantine at institution, alcohol consumption, use of public transport, total amount of offered teaching, and number of hours of paid work.

| | **Absolute scale** | | **Relative scale** | |
|---|---|---|---|---|
| | **Unadjusted** | **Adjusted** | **Unadjusted** | **Adjusted** |
| | | **COVID-19 positive** | | |
| Proportion in-person | -313·3*** | -112·6 | -141·9*** | -51·0 |
| | (-481·3 to -145·2) | (-283·3 to 58·1) | (-218·0 to -65·8) | (-128·3 to 26·3) |
| N | 63 827 | 62 464 | 63 287 | 62 464 |
| | | **COVID-19 tested** | | |
| Proportion in-person | 0·02** | 0·00 | -24·3** | -4·1 |
| | (-0·03 to -0·01) | (-0·02 to 0·01) | (-36·2 to -12·4) | (-17·4 to 9·2) |
| N | 63 827 | 62 464 | 63 827 | 62 464 |
| | | **Quarantine** | | |
| Proportion in-person | -0·02*** | -0·01 | -38·6*** | -14·4 |
| | (-0·03 to -0·01) | (-0·02 to 0·0) | (-53·2 to -24·1) | (-30·1 to 1·8) |
| N | 63 827 | 62 464 | 63 827 | 62 464 |
| | | **Satisfied with teaching** | | |
| Proportion in-person | 1·0*** | 0·6*** | 15·2*** | 9·8*** |
| | (0·9 to 1·1) | (0·6 to 0·7) | (14·0 to 16·5) | (8·4 to 11·1) |
| N | 63 236 | 61 888 | 63 236 | 61 888 |
| | | **Well-being** | | |
| Proportion in-person | 0·6*** | 0·2*** | 9·5*** | 2·6*** |
| | (0·6 to 0·7) | (0·1 to 0·2) | (8·6 to 10·4) | (1·6 to 3·6) |
| N | 63 629 | 62 293 | 63 629 | 62 293 |

*p<0.05, **p< 0.01, ***p<0.001.

Supplementary Figure S1. Impact of actual versus offered proportion of in-person teaching. Panel A visualises results shown in Table 2. Panel B: As Panel A, but outcomes are measured at t-1. Panel C: Fixed effects estimates (see also Table A.1). Panel D: As Panel C, but outcomes are measured at t-1. Panel E: As Panel A, but explanatory variable of interest is actual (self-reported) days on campus. Relative scale is obtained by dividing the coefficient and their standard error on the average of the outcome when the explanatory variable takes zero.

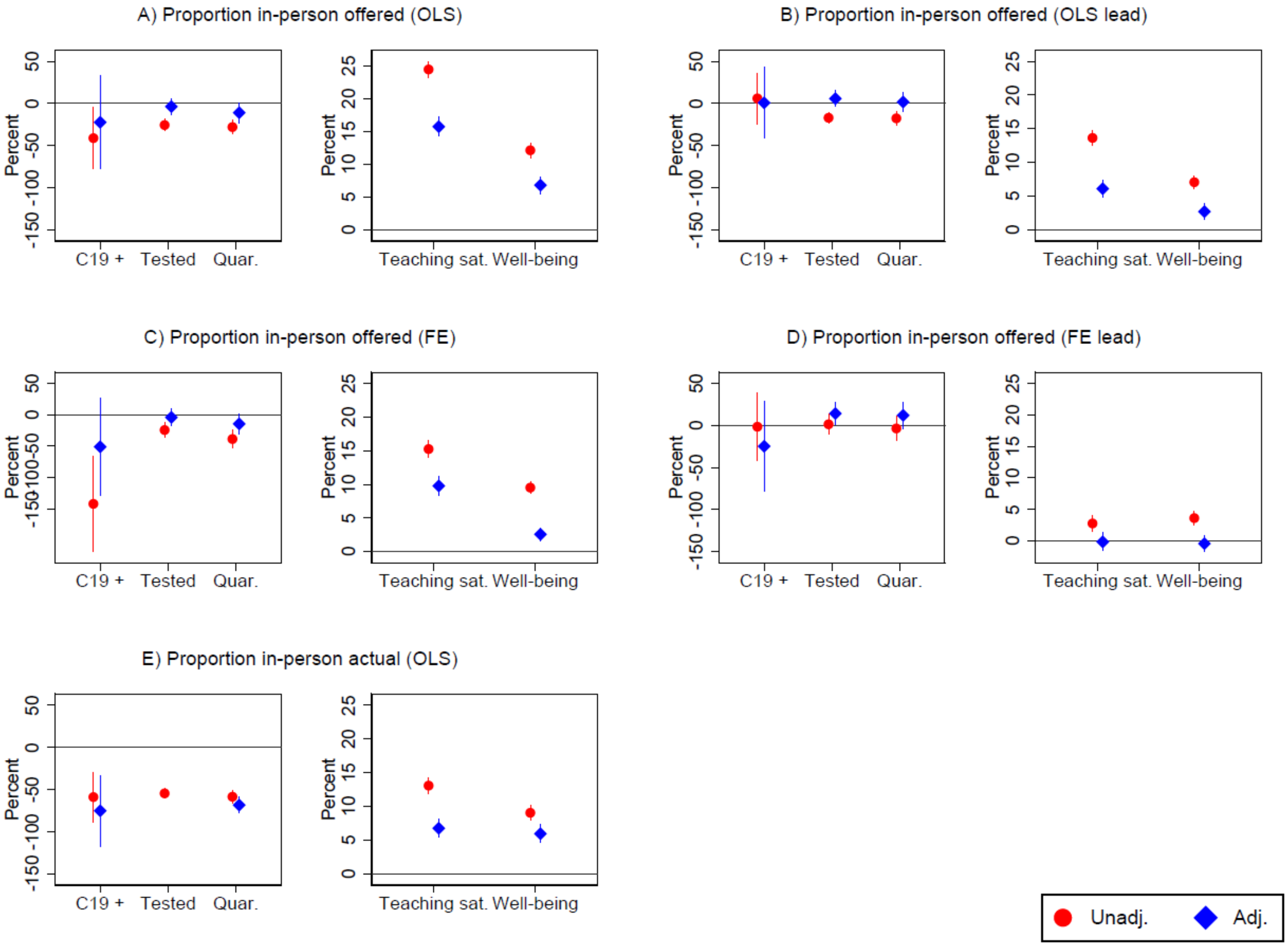

Supplementary Figure S2. Dummy specifications of proportion in-person teaching. Coded 1 for students in the top quartile, otherwise 0 (Panel A) and coded 1 if at least 80%-in person, otherwise 0 (Panel B). Both models are otherwise specified as the main results shown in Table 2. Relative scale is obtained by dividing the coefficient and their standard error on the average of the outcome when the explanatory variable takes zero.

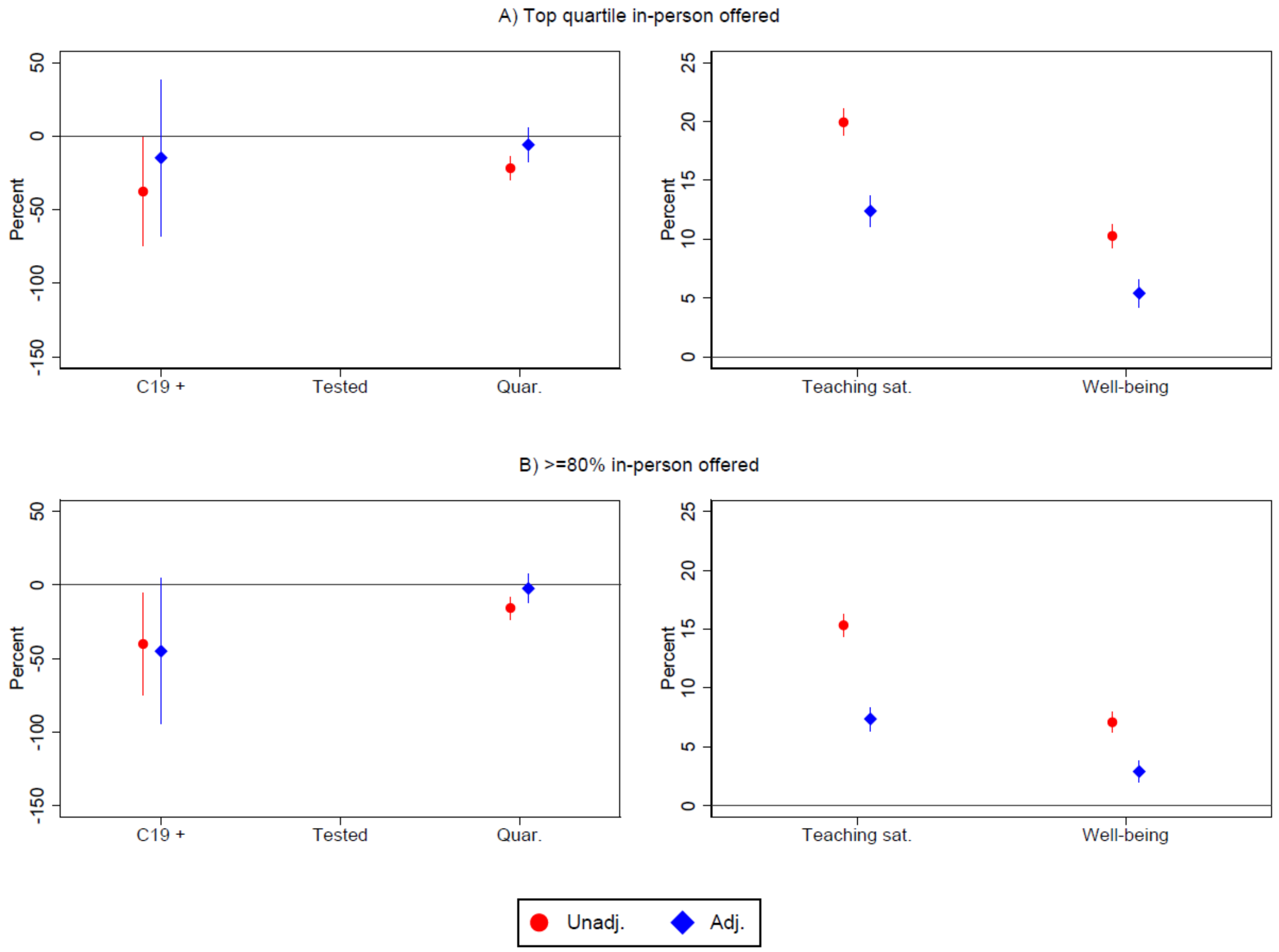

Supplementary Figure S3. Effect of lagged outcomes variable for COVID-19 related outcomes. Note: In this specification, outcomes measured at time t+1 are regressed on predictors measured at time t. Otherwise, the model is identical to the main specification presented in Table 3. Relative scale is obtained by dividing the coefficient and their standard error on the average of the outcome when the explanatory variable takes zero.

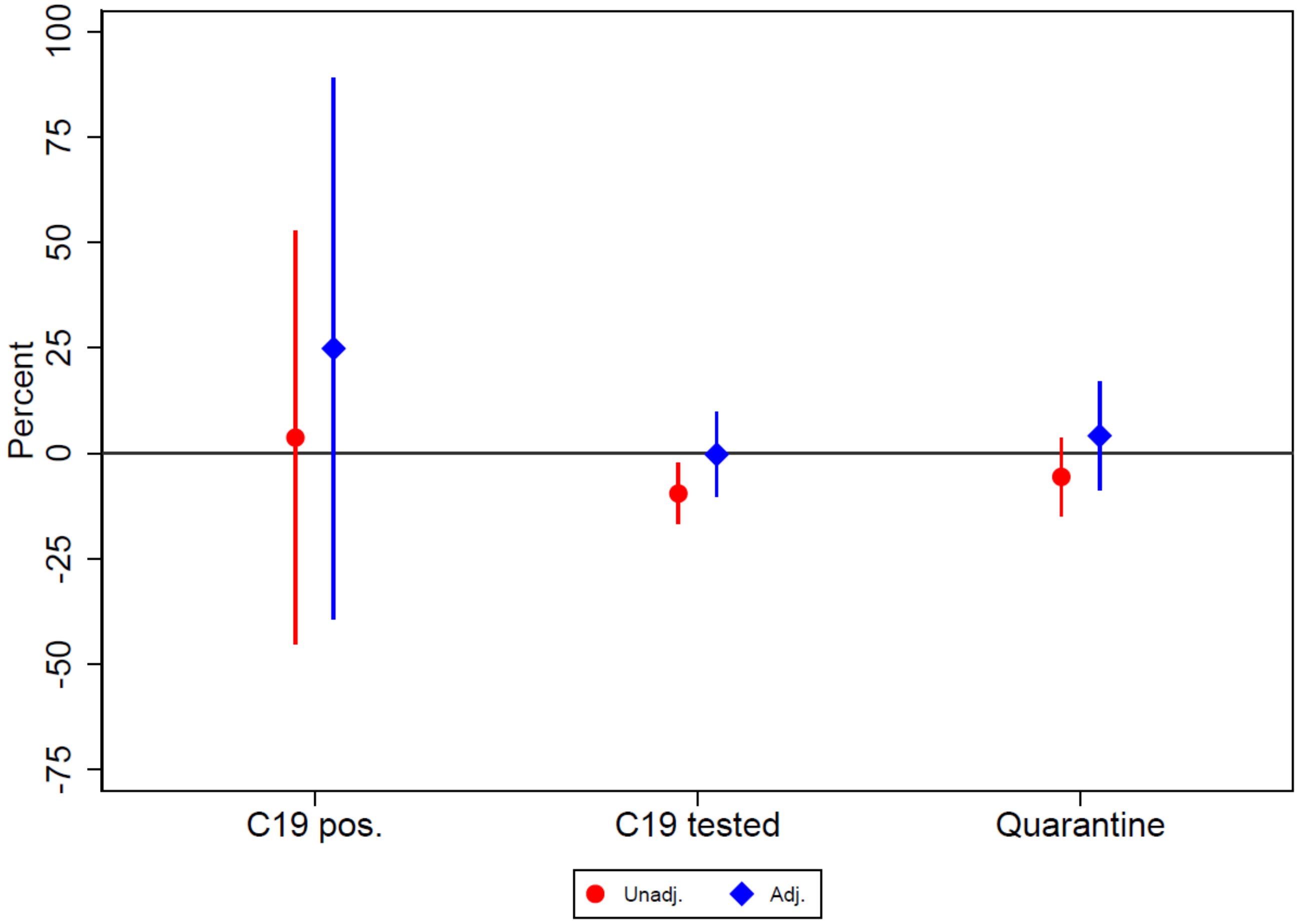
